# A phased approach to unlocking during the COVID-19 pandemic – Lessons from trend analysis

**DOI:** 10.1101/2020.04.20.20072264

**Authors:** Mike Stedman, Mark Davies, Mark Lunt, Arpana Verma, Simon G. Anderson, Adrian H Heald

**Affiliations:** Res Consortium, Andover, Hampshire; Division of Musculoskeletal and Dermatological Sciences; Population Health, Health Services Research and Primary Care, The University of Manchester, and Manchester Academic Health Sciences Centre; University of the West Indies, Cave Hill Campus Barbados; Division of Cardiovascular Sciences, Faculty of Biology, Medicine and Health, The University of Manchester, Manchester, United Kingdom; Department of Diabetes and Endocrinology, Salford; The Faculty of Biology, Medicine and Health and Manchester Academic Health Sciences Centre, University of Manchester, UK

**Keywords:** COVID-19, Unlock, Epidemiology, Strategy

## Abstract

With the COVID-19 pandemic leading to radical political control of social behaviour, including restricted movement outsides homes. Can more detailed analysis of the published confirmed local case data from the pandemic in England using infection ratio and comparing local level data provide a deeper understanding of the wider community infection and inform the future unlocking process.

The historic daily published 78,842 confirmed cases in England up to 13/4/2020 in each of 149 Upper Tier Local Authority (UTLA) were converted to Average Daily Infection Rate (R_ADIR_), an R-value - the number of further people infected by one infected person after their 5-day incubation and during their 5-day infectious phase, and the associated Rate of Change of Infection Rate (ΔIR) also calculated. Results compared to look for significant variances between regions. Stepwise regression was carried out to see what local factors could be linked to the difference in local infection rates.

The peak of COVID-19 infection has passed. The current R_ADIR_ is now below 1. The rate of decline is such that within 14 days it may be below 0.5. There are significant variations in the current RADIR and ΔIR between the UTLAs, suggesting that the disease locally may be at different stages.

Regression analysis across UTLAs found that the only factor that could be related to the fall in RADIR was an increase in the number of confirmed infection/1,000 population. Extrapolation of these results showed that based on assuming a link to increased immunity, unreported community infection may be over 200 times higher than the reported confirmed cases providing evidence that by the end of the second week in April 26% of the population may already have had the disease and so now have increased immunity. Linking these increased estimated infected numbers to recorded deaths indicates a possible mortality rate of 0.14%.

Analysis of the current reported local case data using the infectious ratio does provide greater insight into the current levels of community infection and can be used to make better-informed decisions about the future management of restricted social behaviour and movement

## Background

Severe Acute Respiratory Syndrome Coronavirus-2 (SARS-CoV-2) is the name given to the 2019 novel coronavirus. COVID-19 is the name given to the disease associated with the virus. SARS-CoV-2 is a new strain of coronavirus that has not been previously identified in humans. Following the first recorded cases of SARS-CoV-2 on the 29^th^ January in the United Kingdom, the COVID-19 pandemic has taken a rapidly developing course with a switch by the United Kingdom (UK) Government on the 17^th^ March from a policy of “track and containment” to “mitigation” and initiated social distancing, followed by a comprehensive population lockdown on the 23^rd^ March. The toll on health and lives has been very significant in the UK and elsewhere in the world (1). High-risk groups, based on age and underlying comorbidities, were told to isolate themselves completely for the next 13 weeks (2,3). The rationale was to reduce the impact of the high growth phase of the pandemic on the National Health Service (NHS) with particular focus on Intensive Care Units (ICUs) and High Dependency Units (HDUs) and to keep mortality to a minimum (4,5,6).

Pandemic models forecast that with continuing progress the social lockdown would be relaxed when there is clear evidence of a downturn in infection rates and mortality. There is a trade-off here between balancing the clinical impact of the pandemic with the economic, social and longer-term healthcare impact. This includes considering the impact on diverting resources away from mainstream severe and long-term conditions within primary and secondary care, as well as recognising that the capacity of the population to maintain confinement is limited.

Testing in the very initial phases was carried out on the wider groups who had contacts with diagnosed patients. Testing capacity initiatives have been slow to appear with testing at 5,000/day at the end of March increasing to 10,000/day in April (7). As number of new cases grew and testing capacity limitation was reached testing was restricted to symptomatic hospital-based patients, and more recently as numbers have fallen and testing capacity increased to general practice presentations and NHS staff. Using the total confirmed cases as a sample of the overall levels of population infection is reasonable if the selection rules are consistently applied both over time and geography. While there may be some variations, selection for testing was being restricted during the growth phase and then increased as numbers fall.

This will have first reduced and now increase the numbers of new cases identified. The direction of any error would therefore be to initially to show lower and now relatively higher infection rates.

Given the past community based 3-day doubling infection rate, there are indications that significant part of the population may already have been infected with low grade clinical or subclinical symptoms. This wider non-hospitalised population is likely to continue to grow even with the isolation and social distancing policies.

The ongoing rate of infection is determined to a large extent by the R-value of an infectious disease. The R-value is the number of people infected by one infected person during their infectious phase (8). This value is dependent on the level of local and cross-community social contacts and the proportion of the current population who have not developed immunity through previous exposure. An R-value above 2 suggests more than a doubling of people with the condition during each infectious period and an R-value below 1 is consistent with “suppression” meaning that the virus prevalence will slowly diminish.

The purpose of this paper is to briefly explore data trends from the pandemic in terms of infection rates and policy impact and draw learning points for informing the unlocking process.

## Methods

In England, local government is divided between an Upper Tier (county council) and a Lower Tier (district council). The data of COVID-19 case are published daily for each of the 149 Upper Tier Local Authorities (UTLA) (9) which vary in size from 1.6m to 97k. This study used the latest data download possible.(10)

The UTLA population numbers were taken from GP practice patient numbers published by Lower Layer Super Output Area (LSOA) then aggregated up to their respective UTLA (10,11). The same method was used to aggregate the other population demographic and health characteristics to UTLA level.

### Statistical Analysis

The new cases were calculated and plotted on a timeline with a simple polynomial trend analysis. An exponential curve based on the disease 3 days doubling characteristic linked to the starting data was included for reference.

Two further variables the *Average Daily Infection Rate* (ADIR) and the *Rate of Change of Infection Rate* (ΔIR) were calculated and used to track the national and regional developments in infection rate.

The COVID-19 characteristics incorporated into the analysis are 5 Incubation days and 5 Infectious days. Similar assumptions were made by the Imperial College COVID-19 Response Team in their ‘Impact of non-pharmaceutical interventions (NPIs) to reduce COVID-19 mortality and healthcare demand’ (12). In that paper, the authors stated “We assumed an incubation period of 5.1 days. Infectiousness is assumed to occur from 12 hours before the onset of symptoms for those that are symptomatic and from 4.6 days after infection in those that are asymptomatic with an infectiousness profile over time that results in a 6.5-day mean generation time”.

### Average Daily Infection Rate (R_ADIR_)

The daily infection rate R on any given day is calculated by dividing the infected population i.e. the reported new cases 5 days ahead (corresponding to the incubation period), by the infectious population i.e. an average of new cases over the 5 previous days (corresponding to the infectious period). R_ADIR_ is taken as a rolling average of the R values over the previous 7-days to allow for variation in weekly administrative case count. Therefore

❖ n=Date of Infection
❖ Total Cases (TC)= Daily Reported Total Cumulative Confirmed Cases
❖ New Cases (NC) = TC_(n)_-TC_(n-1)_
❖ Average Infectious group 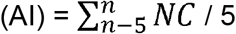 Infectious days
❖ **Infection Ratio (R) = NC** _**(n+5 Incubation Days)**_ **/ AI**_**(n)**_
❖ 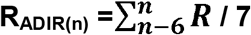 **days in week**

Two sensitivity analyses were considered, first where the condition is faster in incubation and infection (4 days for each) and second where the condition is slower (6 days for each).

### Rate of Change of Infection Rate (ΔIR)

is calculated by taking the slope of the least-squares fit line using the previous 7 days R values.

The relation between the R_ADIR_ and the ΔIR in all the UTLAs was determined and the impact of the disease progression was considered by including the total number of reported cases/,000 population, shown in quartiles.

Local GP practice data taken from various sources were aggregated up to UTLA level. A stepwise regression model linking the UTLA R_ADIR_ to their local community characteristics with weighting by population numbers was carried out with factors included

❖ **Location:** Urban/Rural, Latitude/Longitude, occupants/Household taken from the Office of National Statistics
❖ **Demographics:** % Individual with Age>60, Social Deprivation, % in full-time employment or education, Ethnicity, were taken from NHS GP Practice profiles
❖ **Health:** % population with long term conditions (including hypertension and diabetes), % confident in their own health management were taken from NHS GP Practice Profiles
❖ **COVID-19:** Total reported cases/1,000 population taken from this study 8 April 2020

The regression coefficients for the association between R_ADIR_ and the reported COVID-19 cases/population were used to determine the R_ADIR_ when an UTLA has no reported cases. This is the expected value that the lockdown and increased social distancing delivers on their own on this day. One can also extrapolate to a value of Cases/1,000 pop that would be needed to give a R_ADIR_ = 0 i.e. 100% immunity in the total population. This value can then be used to indicate the relation between reported and community infection levels. Linear extrapolation was used however there may be asymptotic effects that change this number.

The Office of National Statistics has reported a detailed analysis of the total mortality in March associated with COVID-19 (13). This total additional mortality can be related to the total end of March reported cases of COVID-19 which can be uprated by the total potential community infection rate calculated in this report to give an estimate of overall COVID mortality rate.

## Results

As of the 13^th^ April 2020, the number of a confirmed case of COVID-19 in England stands at 69,329 with 10,261 deaths. These covered a total recorded population of 60 million.

Figure 1 shows the reported new cases of COVID-19 have peaked and with reported deaths from COVID-19 which lags new cases by a week also possible approaching a peak.

**Figure 1.**
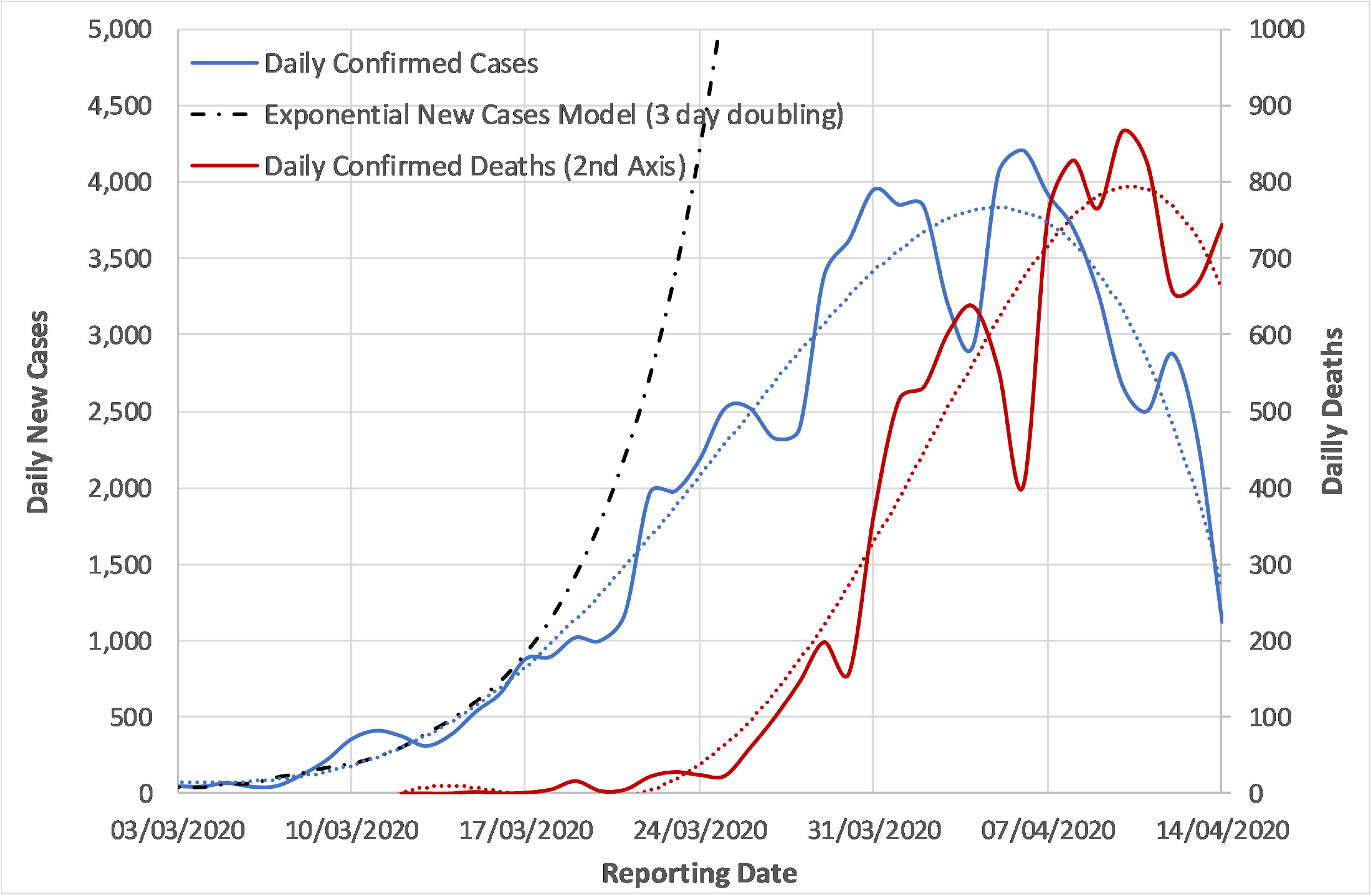
Daily Confirmed New Cases, Model of exponential Case Growth and Confirmed Deaths over the last 6 weeks in England

Analysis for 8^th^ April 2020 (5 days before the latest new cases presentation) shows the national R_ADIR_ was 0.83 with ΔIR at -0.07. In figure 2, the R_ADIR_ is plotted against time, highlighting that the daily R-value breakthrough below 1 has been achieved.

**Figure 2.**
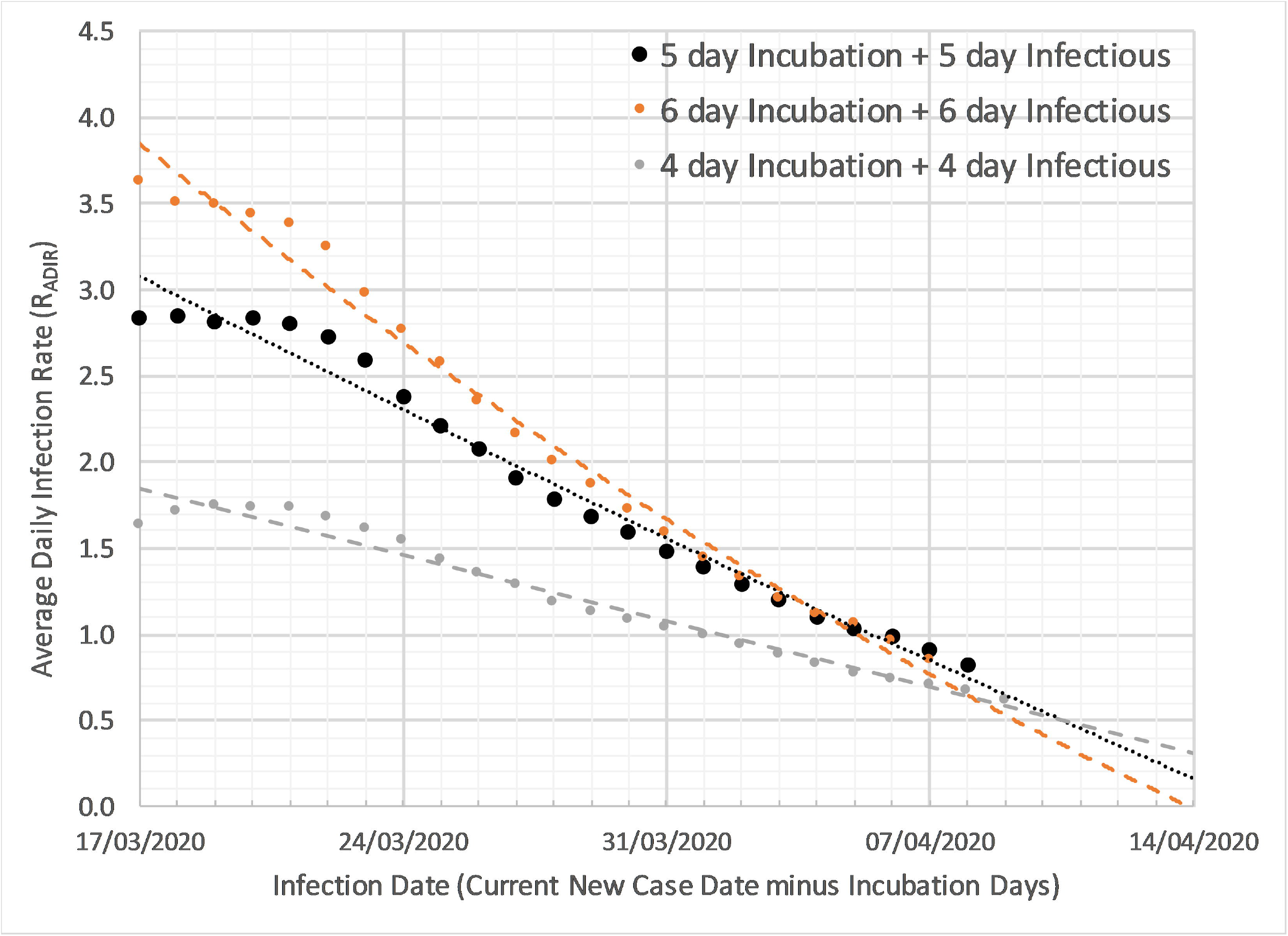
Average Daily Infection Rate (R_ADIR_) for the base model and 2 sensitivity cases

The results of this analysis as shown in Figure 2 show a steady level R_ADIR_ below 3 before the 24/3/2020 lockdown and then a steady fall R_ADIR_ after that. The current infection R_ADIR_ is now below 1 and the rate of decline is such that within 14 days it may be below 0.5. However, given the strong likelihood that the virus will become endemic, a reproductive rate of 0 is unrealistic. Nevertheless, an ongoing R_ADIR_ value well below 1 is a minimum target to reduce rapid re-emergence adopted by the UK Government.

Figure 3 shows the differences in UTLAs of R_ADIR_ and ΔIR. This shows that there is a wide difference between UTLAs with 20 of them still with high infection rates and decreasing more rapidly, while other regions already well below 1 and decreasing more slowly. The inclusion of the number of reported cases/1,000 population quartiles show that those regions with the highest cases/1,000 population now have the lowest infection rates, suggesting there may be a relationship between these two factors.

**Figure 3:**
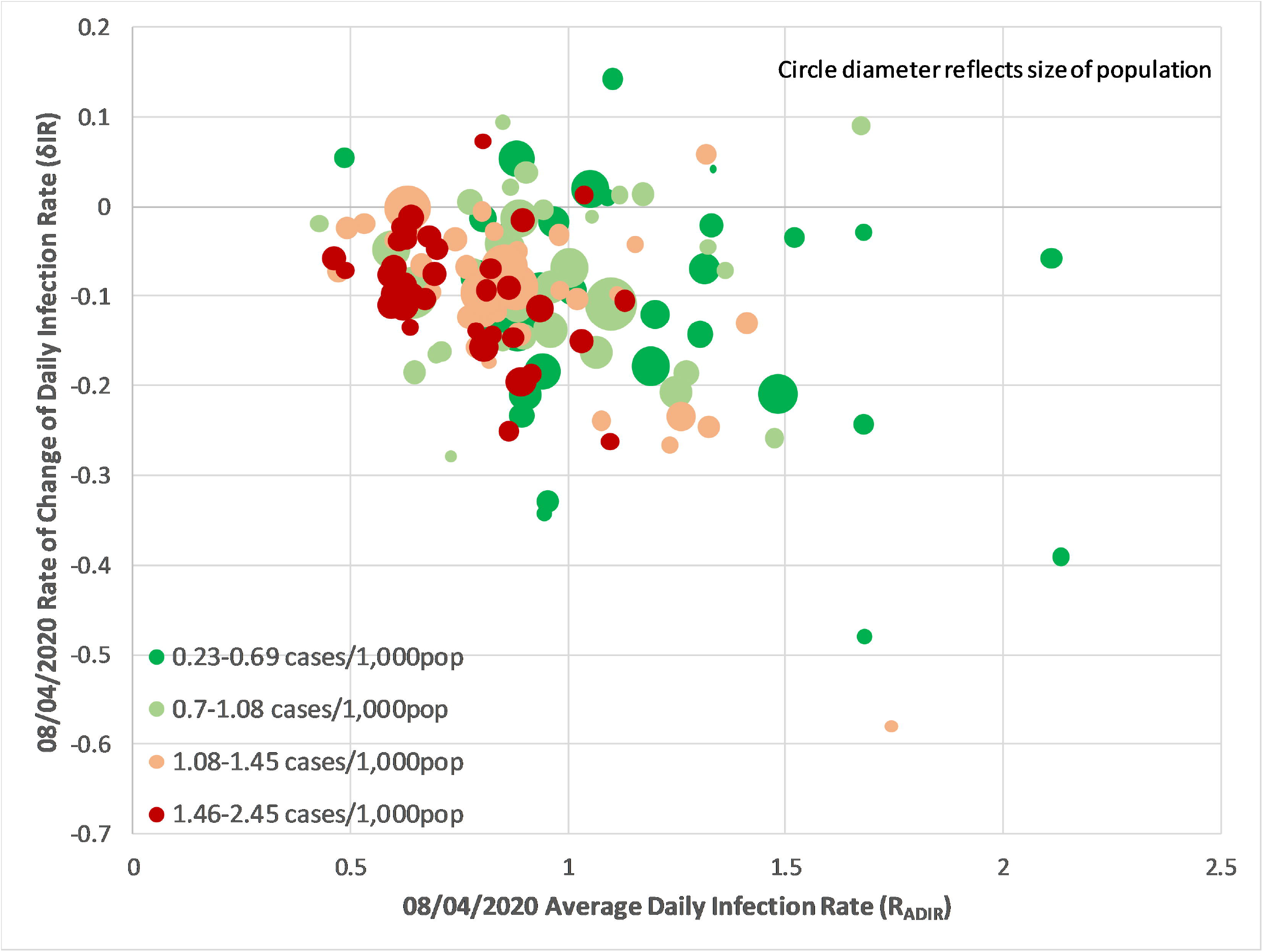
The latest Average Daily Infection Rate (R_ADIR_) versus the Rate of Change of Infection Rate (ΔIR) for each UTLA divided into quartiles for total reported cases/1,000 population

The stepwise regression of the local UTLA factors to R_ADIR_ showed that only one factor total reported cases/1,000 population was significantly linked. In Figure 4, the regression, weighted by local UTLA population, had an r^2^=0.22, p value<0.0001 and the standardised beta of -0.42. Of note here is that the analysis is carried out 5 days before the latest data as, due to the incubation period, that is when the relevant infections would have taken place, and the latest data itself is also subject to ongoing updates.

**Figure 4:**
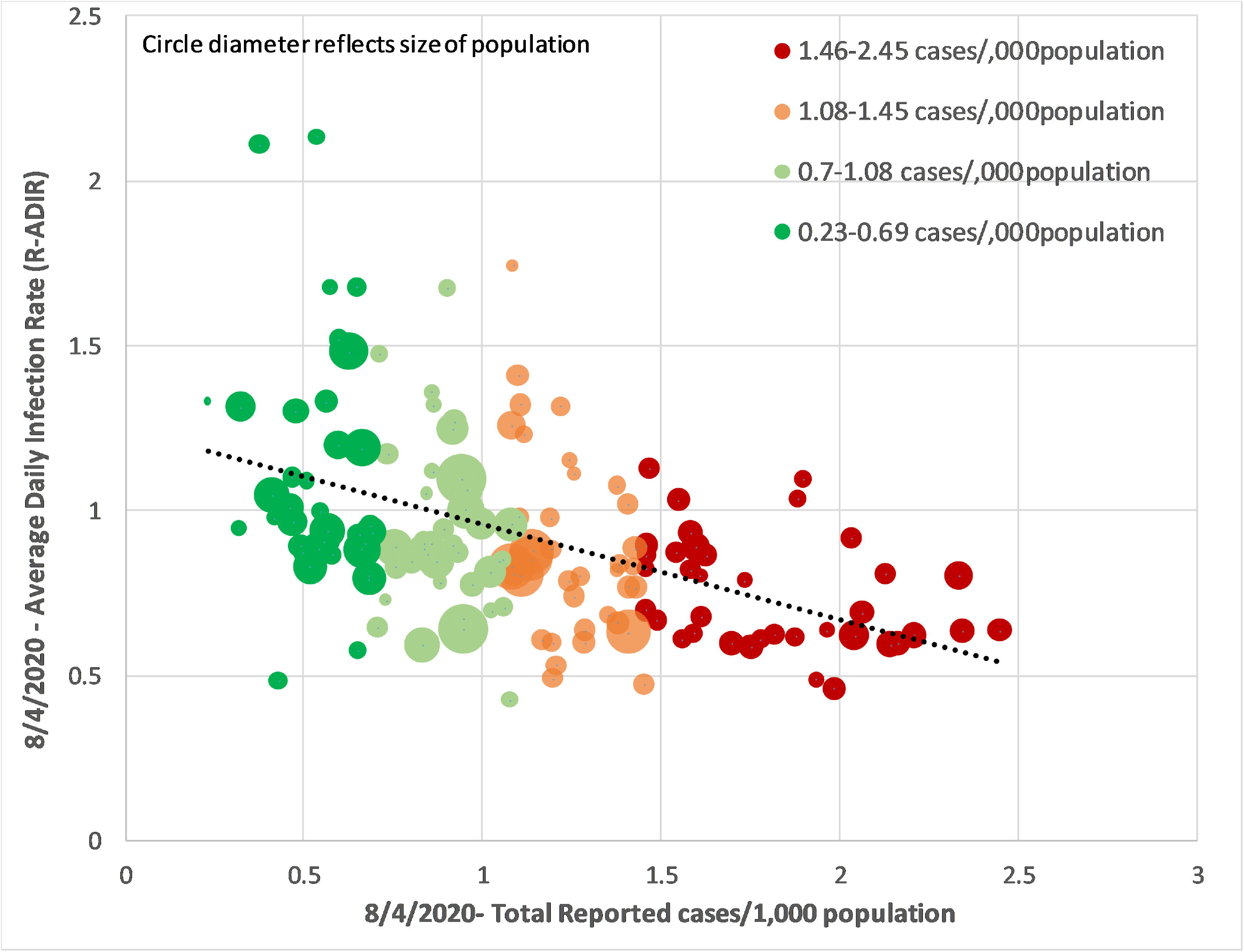
Average Daily Infection Rate (R_ADIR_) for Upper Tier Local Authority linked to Total reported cases/1,000 population

The regression results in an equation R_ADIR_ = 1.20 - 0.26 x Current Total Cases/1,000 population. The reported cases are an unknown fraction of the total community cases. However, one can see that without any reported cases (i.e. no reduced community immunity) a UTLA would have R_ADIR_ of 1.2 - thus the implementation of social distancing has delivered a substantial reduction from the historic R with low number of existing cases at 2.8 (figure 2).

If this relationship is linear then extrapolation (See Figure 4) shows zero R_ADIR_ being achieved at 4.5 reported cases/1,000 community population. Therefore, to achieve full population immunity, this is equivalent to 220 community cases for each reported case. Total reporting of 273,000 confirmed cases would be expected if the total population of 60 million achieved increased immunity. This also suggests with current 73,000 reported cases that 16.1 million (26.8% of the total population) have now been infected.

Applying the 220 difference between community infection and reported cases can also be used to examine mortality. The ONS reported between 1 and 31 March 2020, there was a total of 47,358 deaths. Of these, 3,912 deaths (8%) were reported to have involved the coronavirus (COVID-19)). There were a total 12,288 reported cases of COVID-19 up to 5 days before the end of March; this, according to the above factor (220), is equivalent to a community infection of 2.7 million people. This reflects a mortality rate of 0.14% in the total infected population. If this rate is applied to the total 60 million population then up to 85,000 are at risk of dying.

## Discussion

Having a clear understanding of the historic recovery in the community is a critical piece of information to policymakers as higher levels mitigate the impact associated with relaxing the social constraints. A published piece of work not yet reviewed shows serology results from 1/4/2020 carried out on 3,300 people in Santa Clara California that show 40-80 times as many people in the community have had the disease than was reported by their testing program (14)

The analysis shown in Figure 2 highlights that current lockdown measures are likely to reduce the daily R-value down to negligible levels by the end of April. However, to commence relaxing these measures, we suggest several principles need to be in place to ensure the R-value of COVID-19 does not rise above 1, triggering a second pandemic (there is general acceptance that the disease will inevitably become endemic).

Figure 3 highlights how the disease progression varies across UTLAs and how that impacts the infection rate and its relative speed of change. Regions with history of the most cases/population have the lowest infection rate R_ADIR_ and lowest rate of change in infection rate ΔIR.

Social distancing behaviour and rules implementation could be expected to vary across different communities/groups, and as the different UTLAs have varying amounts of these different communities, examining the variation of infection rate across UTLAs one would hope to see which community groups were responding well and which were responding less well to social distancing. Figure 4 shows the only factor that could be related to the R_ADIR_ in this analysis was the historic number of confirmed number infection/,000 population suggesting that some of the reduction in reported cases is due to the build-up of immunity due to larger numbers of historic cases in the population.

An important comparative R-value reference would be another coronavirus endemic infection, influenza. During seasonal periods, research indicates that influenza has an R-value of around 1.3 (15) and can result in the highest periods up to 200 additional deaths per day above mortality from other causes, although these figures are constrained by the provision of flu vaccine which is available particularly for the high-risk group. However, if the current pandemic can be switched to a similar mortality rate (with carefully phases social behaviour policies in place, along with population testing) then unlocking can be managed in a politically and socially acceptable way. Some observations around this included

a. The principle of self-isolation following infection/symptoms is now well in place in the population
b. The track and contain mechanisms to identify next line contacts of infected people can also be increased with technical support
c. The vulnerable groups can continue to be isolated with their 13-week restriction kept in place but supported by the general population
d. Health service is now better able to cope with the load

Adding to this, experiences with different pandemic policy frameworks suggest that a looser more flexible approach to social activity can be managed if high-risk groups are more carefully protected. This is particularly pertinent given the news this week that elderly care homes are a significant area of both infections and pandemic mortality (16).

The speed of the unlocking process will depend on the level of unlocking. However, what is clear is that with the potentially reduced at-risk population any further peaks will be lower. We looked in UTLAs at the potential determinants of the ADIR and found that the only factor that related to this was the historic number of confirmed number infection/,000 population. This suggests that removing the lockdown from areas with higher historic caseloads should present a lower risk of R-value reversal.

However, a ‘one size fits all’ approach to pandemic policy does not consider the variation in both infection rates and impact across localities. When the data at the regional level is analysed there seems to be a wide variety of R-values and slope of extrapolated R-line over time, implying that unlocking needs to have a certain level of ‘tailoring’ of social behavioural policies and testing to be effective. These differences are likely to be due to differences in local factors such as infection drivers and underlying population morbidities. This has been explored in a separate publication by the same authors (17).

## Conclusion

Unlocking current social restrictions as soon as possible is vital to minimise demand on the economy and the impact of prolonged social containment. However, this must be balanced against containing the current pandemic and minimising future infection waves.

While mindful of the limitations of trend analysis, we believe that several key principles can be derived from the analysis which may aid policy makers in a smoother transition to reducing social containment and sustainably managing the COVID-19 disease. These principles include focusing on achieving low enough R values to keep mortality comparable with influenza, tailoring social behavioural policies to the ongoing tally of latest case numbers and calculating the current R-value within each locality.

We hope this analysis will have relevance and utility for policymakers at national and regional levels in managing the population ‘Unlock’ across the UK and elsewhere.

## Data Availability

All data used for this report is public and published on websites referred to in the references

https://coronavirus.data.gov.uk/

## Figure Legends

Supplementary Table: List of Total Cases and deaths by day and calculated R_ADIR_ and ΔIR

## Disclaimers

Neither Patients nor the public was involved in the design, or conduct, or reporting, or dissemination of our findings.

All authors contributed in relation to the writing of this paper. No author has any conflict of interest.

The manuscript is an honest, accurate, and transparent account of the study being reported. No important aspects of the study have been omitted.

